# A comparative medical genomics approach may facilitate the interpretation of rare missense variation

**DOI:** 10.1101/2023.11.13.23298179

**Authors:** Bushra Haque, George Guirguis, Meredith Curtis, Hera Mohsin, Susan Walker, Michelle M. Morrow, Gregory Costain

## Abstract

**Purpose:** To determine the degree to which likely causal missense variants of single-locus traits in non-human livestock and domestic species have features suggestive of pathogenicity in a human genomic context.

**Methods:** We extracted missense variants from the Online Mendelian Inheritance in Animals database for nine animals (cat, cattle, chicken, dog, goat, horse, pig, rabbit, sheep), mapped coordinates to the human reference genome, and annotated variants using genome analysis tools. We also searched a private commercial laboratory database of genetic testing results from >400,000 individuals with suspected rare disorders.

**Results:** Of 339 variants that were mappable to the same residue and gene in the human genome, 56 had been previously classified with respect to pathogenicity: 31 (55.4%) pathogenic/likely pathogenic, 1 (1.8%) benign/likely benign, and 24 (42.9%) uncertain/other. The odds ratio for a pathogenic/likely pathogenic classification in ClinVar was 7.0 (95% confidence interval: 4.1-12.0, p<0.0001), compared to all other germline missense variants in these same 220 genes. The remaining 283 variants disproportionately had allele frequencies and REVEL scores that supported pathogenicity.

**Conclusion:** Cross-species comparisons could facilitate the interpretation of rare missense variation. These results provide further support for comparative medical genomics approaches that connect big data initiatives in human and veterinary genetics.

## INTRODUCTION

Genome-wide sequencing provides a near-comprehensive view of the exonic single nucleotide variants (SNVs) that contribute to a breadth of human diseases.^1^ Distinguishing pathogenic missense variants that cause rare disease from missense variants that are benign or have no functional consequence remains a major challenge in human genetics.^2^ In the companion field of veterinary medicine, genetic testing of economically important and domesticated animals for Mendelian traits and diseases is increasingly common.^3^ Many first- and second-generation *in silico* prediction tools for missense variants incorporate evolutionary conservation of the amino acid residue,^2 4^ and a new wave of bioinformatic tools are learning from “tolerated” variation in non-human primates and other animals.^5 6^ There is also a long-standing history of isolated observations that specific variants are pathogenic in both human and non-human orthologs (e.g., ^7^). However, the generalizability of a “comparative medical genomics” approach that correlates disease associations of specific variants across species is unknown.

Modeled after Online Mendelian Inheritance in Man (OMIM; omim.org/), the Online Mendelian Inheritance in Animals (OMIA; omia.org/) database includes “likely causal variants” of “single-locus traits” in non-human animal species that were expertly curated from the published literature.^3^ Beginning in 1995, by 2023 OMIA listed 1,577 likely causal variants from 485 species.^3^ Many of the “traits” studied in a veterinary medicine context are severe phenotypes reminiscent of human genetic diseases (e.g., connective tissue, haematologic, skeletal, or neurogenetic disorders; Supplemental Figure 1). We hypothesized that leveraging these easily accessible but previously siloed OMIA data to identify additional evidence for pathogenicity could aid in the interpretation of human missense variants. The purpose of this study was to systematically assess the degree to which pathogenic missense variation observed in a non-human animal genome possess features suggestive of pathogenicity in a human genome context.

## METHODS

### Identification of non-human animal variants and their human equivalents

We extracted all 442 missense single nucleotide variants (SNVs) from OMIA (accessed: January 2023) that were classified as likely causal for Mendelian phenotypes in one of nine animals (cat, cattle, chicken, dog, goat, horse, pig, rabbit, sheep) and where genomic coordinates were reported in OMIA or in the original scientific publication reporting the variant. Four hundred variants were successfully mapped to the human reference genome (GRCh38) using UCSC’s LiftOver tool. We manually inspected each genomic region in the UCSC human genome browser to confirm that (i) the reference amino acid residue in the non-human animal species with the OMIA variant entry matched the reference amino acid residue in humans, and (ii) the alternate amino acid residue would be created through the specific nucleotide change reported in the non-human species; this resulted in the exclusion of an additional 61 variants.

### Searching in public and private databases

We annotated the remaining 339 human genome variants using ANNOVAR^8^ and custom R scripts, including for allele frequency in gnomAD (v3.1.2 and v2.1.1),^9^ presence in ClinVar (download date: August 29, 2022),^10^ phyloP conservation score,^11^ REVEL score,^12^ and AlphaMissense score.^13^ Variants were also searched in Leiden Open Variation Database (LOVD v3.0)^14^ and via Franklin by Genoox (franklin.genoox.com) in winter 2023. We reviewed the primary literature regarding each variant to determine whether the classification or reporting of the variant in one species was directly informed by findings in another species and, for a subset, to summarize the evidence type(s) contributing to a reported association between a specific missense variant and a phenotype in a non-human animal species. Last, we cross-referenced these 339 variants with a large private commercial genetic laboratory (GeneDx) database of genetic testing results from >400,000 individuals with suspected rare Mendelian disorders and their family members. The de-identified data was assessed in accordance with an IRB-approved protocol (WIRB #20171030).

### Statistical methods

Standard descriptive statistics, and parametric and non-parametric tests, were performed using R statistical software, version 4.1.0 (R Foundation for Statistical Computing) with two-tailed statistical significance set at p <0.05.

## RESULTS

In total, 339 missense SNVs across 220 different genes, initially identified in non-human animal species from OMIA, were studied in humans (Figure 1; Supplemental Table 1). There were no variants present in two or more of the non-human species. 159 (72.3%) of the genes were associated with germline Mendelian disease(s) in OMIM (Supplemental Table 2). All but one variant was rare, with a minor allele frequency <0.001 in gnomAD. Most variants (n=228; 67.3%) had REVEL scores ≥0.644 (supporting or greater evidence for pathogenicity) and only 13.6% variants had REVEL scores ≤0.290 (supporting or greater evidence for benign-ness).^15^

**Figure 1.**
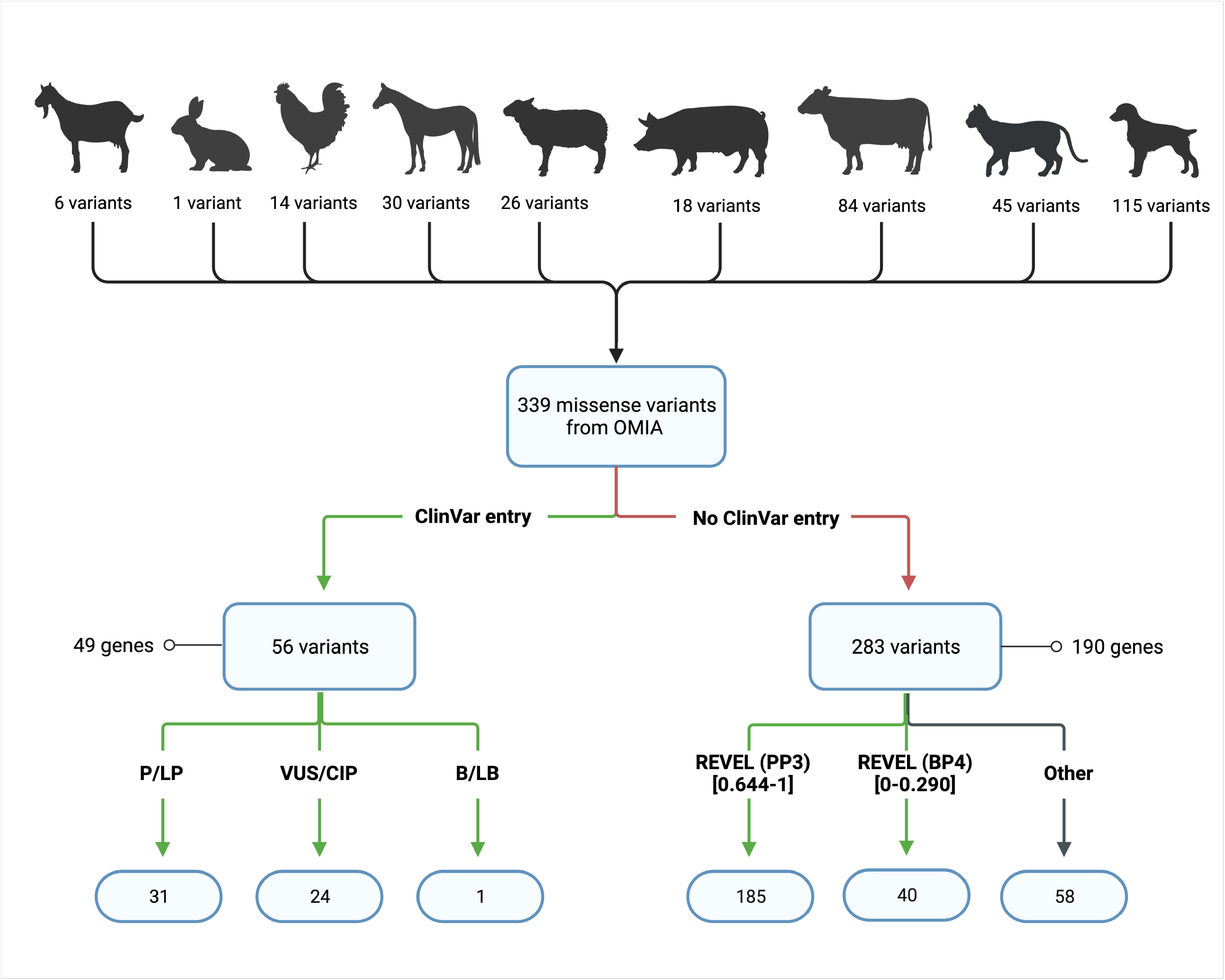
Missense variation from the OMIA database extrapolated to a human genome context. See main text for details. REVEL score thresholds for supporting evidence for pathogenicity (PP3) and for benign-ness (BP4) are from ^15^. B/LB, Benign/Likely benign; P/LP, Pathogenic/Likely pathogenic; VUS/CIP, variant of uncertain significance/conflicting interpretations of pathogenicity. Created with BioRender.com.

### Non-human animal variants were often classified as pathogenic when seen in humans

Of the 339 variants, 56 variants in 49 genes had been previously seen in humans and classified with respect to pathogenicity in ClinVar: 31 (55.4%) as P/LP, 24 (42.9%) as variants of uncertain significance or with conflicting interpretations of pathogenicity (VUS/CIP), and 1 (1.8%) as benign/likely benign (B/LB) (Figure 1). The human Mendelian disease phenotypes associated with these 49 genes were typically concordant with the phenotypes observed in the corresponding non-human animal species (Supplemental Figures 1-2), and included both common (e.g., classic Ehlers-Danlos syndrome) and rare (e.g., geleophysic dysplasia) genetic diseases familiar to medical geneticists. Similarly, 23 of the variants were detected across a total of 172 different families that underwent testing at GeneDx: 15 were classified as P/LP, 7 as VUS, and 1 as B/LB. No additional variants were found in LOVD. The odds ratio for these variants having a P/LP classification in ClinVar was 7.0 (95% confidence interval: 4.1-12.0, p<0.0001), when compared to all other germline missense variants with ClinVar entries in the 220 genes (n=45,925) (Supplemental Figure 3). Determinations of a “likely causal” genotype-phenotype association in a non-human animal species were informed by similar principles as are used in medical genetics practice (Supplemental Figure 4), with an average of 2.75 categories of evidence applied in each report. We found direct references to non-human animal findings informing assessment of the human variant in only 10 cases and noted that the term “OMIA” appears once (accession number VCV000016168.16) in the ClinVar database of 3,373,166 records (search date: June 2023). Conversely, we found references to an equivalent human variant in the initial publication and assessment of the non-human animal variant in 28 instances.

### Understanding discordance between human and non-human variant consequences

We reviewed the variant classified in humans as B/LB, to look for plausible reason(s) for discordant classifications between humans and other animals. The B/LB variant in ClinVar is NM_002386.4(MC1R):c.274G>A;p.(Val92Met) (ClinVar Accession: VCV000014308.11), which has an allele frequency of >0.05 in many human populations. This variant in the melanocyte stimulating hormone receptor gene is a well-studied functional polymorphism associated with pigmentation in humans and in pigs.^16 17^ The impact of the variant is therefore concordant across species, in line with our initial hypothesis. The B/LB variant in the GeneDx dataset is NM_004006.3(DMD):c.5869C>T;p.(Arg1957Trp) and was identified in the hemizygous state in an adult. The REVEL score is 0.22 and hemizygotes (n=3) are reported in the ExAC and ABraOM population databases.^18^ Subsequent review of the original study in pigs^19^ revealed that the variant was associated with “stress syndrome” rather than a muscular dystrophy phenotype, and it remains possible that the variant is a risk factor for similar decompensation in humans.

### In silico scores for the remaining variants were suggestive of pathogenicity in humans

The remaining 283 variants absent from ClinVar were found across 190 genes (Figure 1). Compared with all missense SNVs in ClinVar in these same genes (n=33,526), the REVEL scores of the OMIA variants (median: 0.78; 69.8% ≥0.644) more closely resembled P/LP variants (median: 0.93; 78.4% ≥0.644) than B/LB variants (median: 0.30; 16.2% ≥0.644). To account for potential confounding by residue conservation scores, we restricted to variants with phyloP score ≥0.8. OMIA variants continued to resemble P/LP variants more than B/LB variants with respect to REVEL scores (Figure 2). Findings were similar using AlphaMissense (Supplemental Figure 5).

**Figure 2.**
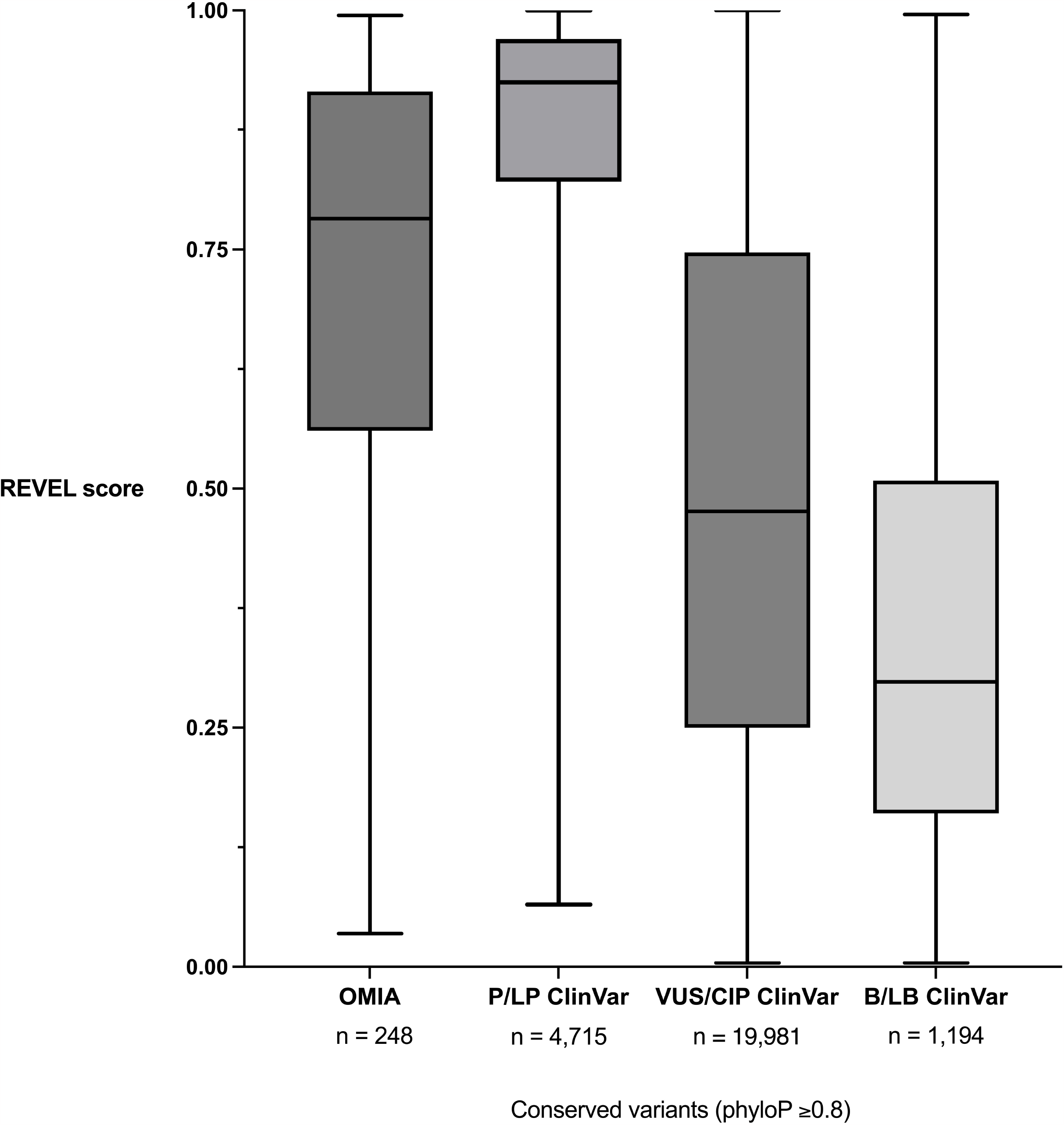
Boxplots of REVEL scores for the OMIA missense variants absent from ClinVar, comparing with other missense variation in ClinVar in the same gene set, restricting all variants to those with phyloP scores of ≥0.8. B/LB, Benign/Likely benign; P/LP, Pathogenic/Likely pathogenic; VUS/CIP, variant of uncertain significance/conflicting interpretations of pathogenicity.

## DISCUSSION

Massive amounts of genomic data are being generated in human populations, but also in domesticated animals like dogs and cattle. While comparative genomics is a well-established field,^20^ and evolutionary conservation of amino acid residues is a foundational component of *in silico* tools for rare variant interpretation, the degree to which the impact of specific rare missense variants at shared residues is concordant across species (“comparative medical genomics”) was understudied. Our results suggest that the missense variants in human genomes that correspond to likely causal variants of single-locus traits in other animals are more likely to be pathogenic (functionally significant). The presence of a variant in OMIA suggests that additional published evidence from a non-human species exists that may be relevant to understanding the impact of the orthologous variant in humans. These observations have implications for rare variant interpretation in medical and veterinary medicine contexts.

While our focus in this report was on the application of non-human animal genomic data to interpreting variation in humans, we suspect that the association is bi-directional. Similarly, although there were no variants present in two or more of the non-human species, we hypothesize that missense variants at conserved residues across two or more species are likely to have a high degree of correlation with respect to functional consequences. There are also several limitations of this study. Consideration of this approach requires conservation of the gene and the residue. There are no consensus variant classification guidelines yet for veterinary medicine / non-human contexts, and the types and quality of evidence supporting a “likely causal” attribution in OMIA will vary by entry. The possibility of circularities in the variant interpretation and reporting process (i.e., that variants were deemed pathogenic in a non-human species and included in OMIA because of pre-existing findings in humans) cannot be excluded. The presence of an orthologous variant in OMIA is suspected but has not been shown to be independent of other lines of evidence typically used in human variant interpretation. There is complete overlap between the ClinVar and GeneDx databases, and so our results should not be overinterpreted as having replicated the findings in two independent cohorts. The added value of the data from GeneDx derives from the relative homogeneity in how evidence was applied for variant interpretation and the ability to look at total number of variant observations/families potentially impacted by OMIA entries.

Our results support the generalizability of a comparative medical genomics approach. The existence of potentially relevant information from a non-human species (as indicated by the variant appearing in OMIA) should prompt a dedicated review of the primary literature supporting the OMIA entry, in situations where additional evidence is needed to interpret the human orthologous variant. The benefits to human medical genetics of leveraging data from veterinary medicine could extend beyond establishing spontaneous large animal disease models for human conditions. Our work is intended to foster new collaborations that bridge previously siloed areas of Mendelian diagnostics, as the many lessons learned from formalizing human DNA variant classifications over the past decade could inform veterinary genetics practice. We anticipate that the coming decade will result in orders of magnitude more genome-wide sequencing in domestic animals and livestock species, through large-scale coordinated academic projects, industry-sponsored research, and direct-to-consumer pet testing. The degree to which the exponential growth of human and veterinary genomic datasets can be integrated and harnessed to improve variant interpretation across both contexts warrants additional study.

## ACKNOWLEDGEMENTS

The authors thank the many contributors to the public and private databases used in this study.

## CONTRIBUTORS

GC and SW conceptualized the study. BH, GG, MC, HM, and MMM curated the data. BH and GG conducted formal analysis of the data. BH and GC acquired the funding for the study. GC supervised the study. BH contributed to all visualization of results. BH, GG, and GC drafted the manuscript, and MC, HM, SW, and MMM were given the opportunity to revise it critically for important intellectual content. All authors give final approval of the submitted version and agree to be accountable for all aspects of the work in ensuring that questions related to the accuracy or integrity of any part of the work are appropriately investigated and resolved.

## PATIENT CONSENT FOR PUBLICATION

Not applicable.

## ETHICS APPROVAL

This secondary use data study was approved by the Research Ethics Board at the Hospital for Sick Children. The de-identified data from GeneDx was assessed in accordance with an IRB-approved protocol (WIRB #20171030).

## DATA AVAILABILITY STATEMENT

All data relevant to the study are available in a public, open access repository (OMIA; omia.org/) and/or are included in the article and uploaded as supplementary information.

