## Supplemental for "A comparative medical genomics approach may facilitate the interpretation of rare missense variation"

**SUPPLEMENTAL FILE FOR:** A comparative medical genomics approach may facilitate the interpretation of rare missense variation

**TABLE OF CONTENTS**

Supplemental Table 1……………………………………………………………………...….…2

Supplemental Table 2………………………………………………………………………......11

Supplemental Figure 1…………………………………………………………...……………..24

Supplemental Figure 2……………………………………………………………...…………..25

Supplemental Figure 3………………………………………………………………...………..26

Supplemental Figure 4…………………………………………………………………...……..27

Supplemental Figure 5…………………………………………………………………...……..28

Supplemental References…………………………………………………………………...…..29

**Supplemental Table 1.** Non-human animal missense variants (n=339) extracted from OMIA and studied in humans. See text for details.

| **Gene** | **Chr** | **Genomic Position (hg19)** | **Reference Allele** | **Alternate Allele** | **Classification of Orthologous Human Variant in ClinVar** |
| --- | --- | --- | --- | --- | --- |
| *ABCA1* | 9 | 107646745 | C | T | ABSENT |
| *ABCA12* | 2 | 215815679 | A | G | ABSENT |
| *ACSL5* | 10 | 114168275 | C | G | ABSENT |
| *ACVR1* | 2 | 158630626 | C | T | LP/P |
| *ADAMTS10* | 19 | 8654389 | C | T | ABSENT |
| *ADAMTS10* | 19 | 8661222 | C | T | ABSENT |
| *ADAMTS17* | 15 | 100673442 | C | T | ABSENT |
| *ADAMTS2* | 5 | 178552032 | C | T | VUS |
| *ADAMTSL2* | 9 | 136406102 | C | T | LP/P |
| *ADAMTSL4* | 1 | 150530585 | G | A | ABSENT |
| *AGTPBP1* | 9 | 88201870 | C | G | ABSENT |
| *AGXT* | 2 | 241808659 | G | A | LP/P |
| *AGXT* | 2 | 241810860 | G | A | ABSENT |
| *ALDH5A1* | 6 | 24523122 | G | A | ABSENT |
| *ALMS1* | 2 | 73777489 | G | C | ABSENT |
| *ALPL* | 1 | 21903126 | T | G | ABSENT |
| *AR* | X | 66931514 | G | C | ABSENT |
| *AR* | X | 66937392 | C | T | ABSENT |
| *ARSB* | 5 | 78076270 | C | T | VUS |
| *ARSB* | 5 | 78076401 | A | G | ABSENT |
| *ARSB* | 5 | 78181645 | C | T | ABSENT |
| *ARSG* | 17 | 66339822 | G | A | VUS |
| *ASIP* | 20 | 32856863 | C | T | ABSENT |
| *ASIP* | 20 | 32856947 | T | A | ABSENT |
| *ASPA* | 17 | 3402296 | G | C | ABSENT |
| *ASPRV1* | 2 | 70187847 | A | G | ABSENT |
| *ATF2* | 2 | 175986219 | A | C | ABSENT |
| *ATG4D* | 19 | 10663609 | G | A | ABSENT |
| *ATP13A2* | 1 | 17323577 | G | A | ABSENT |
| *ATP2A1* | 16 | 28909686 | C | T | ABSENT |
| *ATP2A1* | 16 | 28898972 | G | T | ABSENT |
| *ATP2A1* | 16 | 28898747 | G | T | ABSENT |
| *ATP7A* | X | 77245098 | C | T | ABSENT |
| *BCKDHA* | 19 | 41930425 | C | T | ABSENT |
| *BEST1* | 11 | 61724316 | G | A | ABSENT |
| *BMP15* | X | 50659525 | G | T | ABSENT |
| *BMP15* | X | 50659434 | A | C | ABSENT |
| *BMP15* | X | 50659387 | G | A | ABSENT |
| *BMP15* | X | 50659375 | C | T | ABSENT |
| *BMP3* | 4 | 81974618 | C | A | ABSENT |
| *BMPR1B* | 4 | 96051173 | A | G | ABSENT |
| *BRAF* | 7 | 140453136 | A | T | LP/P |
| *CAD* | 2 | 27447710 | A | G | VUS |
| *CAPN1* | 11 | 64950951 | G | A | ABSENT |
| *CAT* | 11 | 34478287 | G | A | ABSENT |
| *CD320* | 19 | 8368785 | G | C | ABSENT |
| *CDH23* | 10 | 73330622 | C | T | ABSENT |
| *CHAT* | 10 | 50827789 | G | A | CIP |
| *CHST6* | 16 | 75512976 | G | T | ABSENT |
| *CLCN1* | 7 | 143021535 | C | G | ABSENT |
| *CLCN1* | 7 | 143048744 | G | C | ABSENT |
| *CLCN1* | 7 | 143039214 | A | C | ABSENT |
| *CLCN1* | 7 | 143016944 | G | A | ABSENT |
| *CLN6* | 15 | 68500588 | A | G | CIP |
| *CLN6* | 15 | 68510888 | G | A | ABSENT |
| *CNGA3* | 2 | 99012861 | C | T | LP/P |
| *CNGA3* | X | 150912464 | G | A | ABSENT |
| *CNGB3* | 8 | 87679206 | C | T | ABSENT |
| *CNGB3* | 8 | 87679206 | C | T | ABSENT |
| *CNTNAP1* | 17 | 40845375 | G | A | ABSENT |
| *COL10A1* | 6 | 116441496 | C | T | ABSENT |
| *COL11A2* | 6 | 33157186 | C | G | ABSENT |
| *COL1A1* | 17 | 48273017 | C | T | ABSENT |
| *COL1A1* | 17 | 48263763 | A | T | ABSENT |
| *COL1A1* | 17 | 48272838 | C | G | ABSENT |
| *COL2A1* | 12 | 48378812 | C | T | ABSENT |
| *COL2A1* | 12 | 48376666 | C | T | ABSENT |
| *COL2A1* | 12 | 48372397 | C | T | ABSENT |
| *COL2A1* | 12 | 48372091 | C | T | ABSENT |
| *COL2A1* | 12 | 48371210 | C | T | ABSENT |
| *COL5A1* | 9 | 137707826 | G | A | VUS |
| *COL5A2* | 2 | 189921724 | C | A | ABSENT |
| *COL6A3* | 2 | 238274501 | G | A | VUS |
| *COL7A1* | 3 | 48613963 | C | T | ABSENT |
| *COLQ* | 3 | 15497411 | C | T | LP/P |
| *COLQ* | 3 | 15498031 | A | G | ABSENT |
| *COLQ* | 3 | 15499767 | C | T | ABSENT |
| *COPA* | 1 | 160302256 | G | A | ABSENT |
| *CORIN* | 4 | 47625754 | G | A | ABSENT |
| *CORIN* | 4 | 47695070 | C | T | ABSENT |
| *CPT1C* | 19 | 50200599 | G | A | ABSENT |
| *CSNK1G2* | 19 | 1978903 | G | C | ABSENT |
| *CTSD* | 11 | 1778655 | C | T | ABSENT |
| *CTSD* | 11 | 1775313 | C | T | ABSENT |
| *CYB5R3* | 22 | 43023363 | T | G | ABSENT |
| *CYB5R3* | 22 | 43027384 | C | T | ABSENT |
| *CYP26C1* | 10 | 94822610 | T | C | ABSENT |
| *DKK4* | 8 | 42233272 | C | T | ABSENT |
| *DKK4* | 8 | 42234511 | G | A | ABSENT |
| *DMD* | X | 32360270 | G | A | CIP |
| *DNM1* | 9 | 130982538 | G | T | ABSENT |
| *DNM2* | 19 | 10909219 | C | T | LP/P |
| *DPYS* | 8 | 105405152 | C | T | LP/P |
| *DSP* | 6 | 7584358 | C | A | ABSENT |
| *DUOX2* | 15 | 45401730 | T | C | ABSENT |
| *EDN2* | 1 | 41949787 | C | T | ABSENT |
| *EDNRA* | 4 | 148453825 | G | T | ABSENT |
| *EDNRA* | 4 | 148461045 | G | A | ABSENT |
| *ENAM* | 4 | 71507856 | C | T | ABSENT |
| *ENTREP2* | 15 | 29428645 | T | C | ABSENT |
| *ETFDH* | 4 | 159616656 | T | G | ABSENT |
| *F11* | 4 | 187207634 | G | A | VUS |
| *F12* | 5 | 176829582 | C | G | ABSENT |
| *F8* | X | 154091390 | A | T | ABSENT |
| *F8* | X | 154189400 | G | C | ABSENT |
| *F8* | X | 154185266 | C | T | ABSENT |
| *F9* | X | 138644124 | G | A | ABSENT |
| *FAM20C* | 7 | 295908 | C | T | ABSENT |
| *FAM83G* | 17 | 18907200 | C | G | ABSENT |
| *FBN1* | 15 | 48777685 | C | T | ABSENT |
| *FGF5* | 4 | 81207590 | G | A | ABSENT |
| *FGF5* | 4 | 81207488 | A | C | ABSENT |
| *FGF5* | 4 | 81188256 | G | T | ABSENT |
| *FGF5* | 4 | 81207591 | C | T | ABSENT |
| *FGFR2* | 10 | 123279562 | C | A | LP/P |
| *FGFR3* | 4 | 1808341 | T | A | ABSENT |
| *FLCN* | 17 | 17125830 | T | C | ABSENT |
| *FZD7* | 2 | 202900611 | G | C | ABSENT |
| *G6PC1* | 17 | 41059562 | G | C | ABSENT |
| *GALC* | 14 | 88450799 | T | G | ABSENT |
| *GART* | 21 | 34900853 | T | G | ABSENT |
| *GDF9* | 5 | 132199997 | G | A | LP/P |
| *GDF9* | 5 | 132197364 | T | G | ABSENT |
| *GDF9* | 5 | 132197459 | G | A | ABSENT |
| *GDF9* | 5 | 132197532 | C | T | ABSENT |
| *GDF9* | 5 | 132197609 | A | C | ABSENT |
| *GDF9* | 5 | 132197700 | G | A | ABSENT |
| *GFAP* | 17 | 42990701 | C | T | LP/P |
| *GLB1* | 3 | 33058235 | C | G | LP/P |
| *GLB1* | 3 | 33114105 | C | T | ABSENT |
| *GLB1* | 3 | 33099625 | C | A | ABSENT |
| *GUSB* | 7 | 65435319 | A | C | CIP |
| *GUSB* | 7 | 65435316 | G | A | ABSENT |
| *GUSB* | 7 | 65439917 | C | T | ABSENT |
| *GUSB* | 7 | 65441045 | G | A | ABSENT |
| *HCRTR2* | 6 | 55039545 | G | A | ABSENT |
| *HES7* | 17 | 8027398 | A | G | ABSENT |
| *HEXA* | 15 | 72641439 | C | T | LP/P |
| *HEXA* | 15 | 72637818 | G | A | ABSENT |
| *HMBS* | 11 | 118959966 | G | A | ABSENT |
| *IARS1* | 9 | 95050449 | C | G | ABSENT |
| *IFT122* | 3 | 129234353 | G | A | ABSENT |
| *ITGA2B* | 17 | 42460879 | C | G | ABSENT |
| *ITGA3* | 17 | 48148681 | C | T | ABSENT |
| *ITGB2* | 21 | 46323396 | T | C | ABSENT |
| *ITGB2* | 21 | 46330239 | C | G | ABSENT |
| *KCNG1* | 20 | 49620870 | C | A | ABSENT |
| *KCNJ10* | 1 | 160011337 | A | G | ABSENT |
| *KDM2B* | 12 | 121880815 | C | T | ABSENT |
| *KDSR* | 18 | 61018135 | C | T | ABSENT |
| *KIF1C* | 17 | 4905937 | G | A | ABSENT |
| *KIF3B* | 20 | 30898580 | G | A | ABSENT |
| *KIT* | 4 | 55604682 | G | A | VUS |
| *KIT* | 4 | 55595543 | T | C | VUS |
| *KIT* | 4 | 55595543 | T | A | ABSENT |
| *KIT* | 4 | 55595523 | A | T | ABSENT |
| *KIT* | 4 | 55594269 | G | A | ABSENT |
| *KIT* | 4 | 55594031 | C | T | ABSENT |
| *KIT* | 4 | 55589840 | A | G | ABSENT |
| *KIT* | 4 | 55569989 | G | A | ABSENT |
| *KIT* | 4 | 55565844 | T | C | ABSENT |
| *KIT* | 4 | 55598160 | A | T | ABSENT |
| *KIT* | 4 | 55575645 | A | G | ABSENT |
| *KIT* | 4 | 55599304 | T | A | ABSENT |
| *KLKB1* | 4 | 187172760 | T | A | ABSENT |
| *KRT25* | 17 | 38911258 | C | T | ABSENT |
| *KRT27* | 17 | 38938470 | G | C | ABSENT |
| *KRT5* | 12 | 52910431 | C | T | LP/P |
| *KRT71* | 12 | 52944024 | G | A | ABSENT |
| *L2HGDH* | 14 | 50713867 | T | C | ABSENT |
| *LAMA3* | 18 | 21512141 | A | T | ABSENT |
| *LAMB3* | 1 | 209801494 | A | G | ABSENT |
| *LAMP3* | 3 | 182841997 | C | T | ABSENT |
| *LDLR* | 19 | 11215925 | C | T | LP/P |
| *LOXHD1* | 18 | 44085947 | C | G | ABSENT |
| *LRP4* | 11 | 46897459 | C | T | ABSENT |
| *LRP4* | 11 | 46903348 | C | T | ABSENT |
| *LVRN* | 5 | 115298993 | G | A | ABSENT |
| *LYST* | 1 | 235929441 | T | C | ABSENT |
| *MAN2B1* | 19 | 12774621 | C | T | ABSENT |
| *MAN2B1* | 19 | 12772142 | A | G | ABSENT |
| *MARS2* | 2 | 198571682 | G | A | ABSENT |
| *MC1R* | 16 | 89985916 | G | A | VUS |
| *MC1R* | 16 | 89986316 | A | C | VUS |
| *MC1R* | 16 | 89986070 | T | A | LB/B |
| *MC1R* | 16 | 89985884 | T | C | ABSENT |
| *MC1R* | 16 | 89985946 | G | A | ABSENT |
| *MC1R* | 16 | 89985962 | T | C | ABSENT |
| *MC1R* | 16 | 89985929 | G | A | ABSENT |
| *MC1R* | 16 | 89986090 | C | T | ABSENT |
| *MC1R* | 16 | 89986467 | C | G | ABSENT |
| *MC1R* | 16 | 89985916 | G | A | ABSENT |
| *MC1R* | 16 | 89986148 | C | T | ABSENT |
| *MC1R* | 16 | 89986027 | G | A | ABSENT |
| *MC1R* | 16 | 89985962 | T | C | ABSENT |
| *MC1R* | 16 | 89985940 | G | A | ABSENT |
| *MC1R* | 16 | 89986384 | G | A | ABSENT |
| *MC1R* | 16 | 89985884 | T | A | ABSENT |
| *MC1R* | 16 | 89986027 | G | A | ABSENT |
| *MC4R* | 18 | 58038691 | C | T | ABSENT |
| *MITF* | 3 | 70005618 | G | T | VUS |
| *MITF* | 3 | 70008444 | G | A | CIP |
| *MITF* | 3 | 70001032 | A | G | LP/P |
| *MITF* | 3 | 70008453 | T | C | ABSENT |
| *MLPH* | 2 | 238402172 | C | T | LP/P |
| *MLPH* | 2 | 238428672 | G | C | ABSENT |
| *MOCOS* | 18 | 33767639 | T | C | ABSENT |
| *MRC2* | 17 | 60754698 | T | G | ABSENT |
| *MSTN* | 2 | 190927132 | A | G | ABSENT |
| *MSTN* | 2 | 190927009 | G | C | ABSENT |
| *MSTN* | 2 | 190924991 | C | T | ABSENT |
| *MSTN* | 2 | 190922174 | C | T | ABSENT |
| *MTBP* | 8 | 121534921 | G | A | ABSENT |
| *MTM1* | X | 149807426 | C | T | ABSENT |
| *MTM1* | X | 149826391 | A | C | ABSENT |
| *MX1* | 21 | 42830462 | G | A | ABSENT |
| *MYBPC1* | 12 | 102036319 | T | G | LP/P |
| *MYBPC3* | 11 | 47372991 | C | G | CIP |
| *MYBPC3* | 11 | 47359086 | G | A | ABSENT |
| *MYH1* | 17 | 10416047 | T | C | ABSENT |
| *MYH7* | 14 | 23883224 | C | T | VUS |
| *MYO7A* | 11 | 76901153 | G | A | LP/P |
| *NAGLU* | 17 | 40695378 | G | A | LP/P |
| *NAPEPLD* | 7 | 102760427 | C | G | ABSENT |
| *NDRG1* | 8 | 134274323 | C | A | ABSENT |
| *NECAP1* | 12 | 8245519 | G | A | ABSENT |
| *NHLRC2* | 10 | 115644032 | T | C | ABSENT |
| *NPC1* | 18 | 21119363 | C | G | ABSENT |
| *NPC1* | 18 | 21136211 | T | G | ABSENT |
| *NPC1* | 18 | 21118578 | G | C | ABSENT |
| *NSDHL* | X | 152031158 | A | G | ABSENT |
| *NSDHL* | X | 152036164 | G | A | ABSENT |
| *NUBPL* | 14 | 32142695 | C | A | ABSENT |
| *P3H2* | 3 | 189688652 | C | G | ABSENT |
| *PAX3* | 2 | 223161809 | C | T | ABSENT |
| *PAX3* | 2 | 223161923 | G | C | ABSENT |
| *PCDH15* | 10 | 55780104 | G | C | ABSENT |
| *PFAS* | 17 | 8172081 | C | T | ABSENT |
| *PFKM* | 12 | 48527220 | C | T | ABSENT |
| *PIGN* | 18 | 59824406 | G | A | ABSENT |
| *PITX3* | 10 | 103990842 | C | G | ABSENT |
| *PKD1* | 16 | 2150031 | C | T | ABSENT |
| *PKLR* | 1 | 155264390 | A | G | LP/P |
| *PKLR* | 1 | 155264148 | C | T | ABSENT |
| *PLN* | 6 | 118880110 | G | A | CIP |
| *PLOD1* | 1 | 12034713 | G | A | CIP |
| *PLP1* | X | 103040616 | A | C | ABSENT |
| *PLP2* | X | 49028397 | C | T | ABSENT |
| *PMEL* | 12 | 56359732 | C | T | ABSENT |
| *PNKP* | 19 | 50370313 | T | C | ABSENT |
| *PNPLA8* | 7 | 108128384 | C | T | ABSENT |
| *PPARD* | 6 | 35378959 | G | A | ABSENT |
| *PPIB* | 15 | 64455071 | C | T | ABSENT |
| *PRCD* | 17 | 74536228 | G | A | LP/P |
| *PRKAG3* | 2 | 219693283 | C | T | VUS |
| *PRL* | 6 | 22287660 | A | C | ABSENT |
| *PRNP* | 20 | 4680464 | G | A | LP/P |
| *PRNP* | 20 | 4680285 | A | G | LP/P |
| *PRNP* | 20 | 4680245 | G | A | ABSENT |
| *PRNP* | 20 | 4680283 | A | G | ABSENT |
| *PRNP* | 20 | 4680318 | G | A | ABSENT |
| *PRNP* | 20 | 4680264 | C | T | ABSENT |
| *PRNP* | 20 | 4680318 | G | A | ABSENT |
| *PSMB7* | 9 | 127177131 | A | C | ABSENT |
| *PYGM* | 11 | 64520595 | G | A | ABSENT |
| *RAB24* | 5 | 176730177 | T | G | ABSENT |
| *RABGGTB* | 1 | 76257870 | A | G | ABSENT |
| *RASGRP2* | 11 | 64506944 | A | G | ABSENT |
| *RDH5* | 12 | 56115704 | G | T | ABSENT |
| *RETREG1* | 5 | 16479075 | G | A | ABSENT |
| *RHO* | 3 | 129247587 | C | G | ABSENT |
| *RYR1* | 19 | 38946154 | T | C | LP/P |
| *RYR1* | 19 | 38991282 | C | G | ABSENT |
| *RYR1* | 19 | 38948185 | C | T | ABSENT |
| *SCN8A* | 12 | 52200168 | G | T | ABSENT |
| *SEL1L* | 14 | 81950643 | A | G | ABSENT |
| *SERPINH1* | 11 | 75282848 | T | C | ABSENT |
| *SGSH* | 17 | 78188504 | G | A | CIP |
| *SLC12A1* | 15 | 48527101 | C | T | ABSENT |
| *SLC24A5* | 15 | 48414204 | T | A | ABSENT |
| *SLC25A12* | 2 | 172669974 | A | G | ABSENT |
| *SLC25A46* | 5 | 110079477 | C | T | ABSENT |
| *SLC2A9* | 4 | 9982268 | C | A | ABSENT |
| *SLC35A3* | 1 | 100476990 | G | T | ABSENT |
| *SLC35A3* | 1 | 100459183 | C | A | ABSENT |
| *SLC36A1* | 5 | 150843158 | C | G | ABSENT |
| *SLC39A4* | 8 | 145639765 | C | G | ABSENT |
| *SLC3A1* | 2 | 44539746 | C | T | LP/P |
| *SLC45A2* | 5 | 33954504 | G | T | ABSENT |
| *SLC45A2* | 5 | 33963887 | T | C | ABSENT |
| *SLC45A2* | 5 | 33944868 | C | T | ABSENT |
| *SLC45A2* | 5 | 33984384 | C | T | ABSENT |
| *SLC45A2* | 5 | 33982446 | C | T | ABSENT |
| *SLC45A2* | 5 | 33954510 | C | T | ABSENT |
| *SLC5A3* | 21 | 35468849 | C | T | ABSENT |
| *SLC6A5* | 11 | 20628679 | T | C | ABSENT |
| *SLC7A9* | 19 | 33353031 | C | T | LP/P |
| *SLC7A9* | 19 | 33350748 | A | T | ABSENT |
| *SLC7A9* | 19 | 33333132 | G | A | ABSENT |
| *SLC7A9* | 19 | 33349368 | C | T | ABSENT |
| *SMC2* | 9 | 106900433 | T | C | ABSENT |
| *SOD1* | 21 | 33036151 | G | A | ABSENT |
| *SOWAHB* | 4 | 77817829 | G | T | ABSENT |
| *SPAST* | 2 | 32370074 | G | A | LP/P |
| *STAT5B* | 17 | 40359728 | T | G | ABSENT |
| *SUGT1* | 13 | 53261902 | T | C | ABSENT |
| *SUV39H2* | 10 | 14941660 | T | G | ABSENT |
| *TBXT* | 6 | 166580884 | T | C | ABSENT |
| *TBXT* | 6 | 166580891 | G | C | ABSENT |
| *TECPR2* | 14 | 102963981 | C | T | VUS |
| *TNXB* | 6 | 32053766 | C | T | ABSENT |
| *TPO* | 2 | 1440068 | G | A | ABSENT |
| *TPO* | 2 | 1491745 | C | T | ABSENT |
| *TRPV4* | 12 | 110236547 | C | A | LP/P |
| *TSEN54* | 17 | 73513639 | G | A | ABSENT |
| *TUBB1* | 20 | 57594582 | G | A | ABSENT |
| *TUBB1* | 20 | 57599227 | G | A | ABSENT |
| *TUBD1* | 17 | 57955604 | T | C | ABSENT |
| *TUBGCP5* | 15 | 22840245 | C | A | ABSENT |
| *TYR* | 11 | 88924454 | G | A | LP/P |
| *TYR* | 11 | 88911351 | G | A | LP/P |
| *TYR* | 11 | 88961072 | C | A | ABSENT |
| *TYRP1* | 9 | 12695772 | C | A | ABSENT |
| *TYRP1* | 9 | 12708035 | C | T | ABSENT |
| *TYRP1* | 9 | 12694117 | T | A | ABSENT |
| *TYRP1* | 9 | 12694121 | G | A | ABSENT |
| *TYRP1* | 9 | 12702382 | T | G | ABSENT |
| *TYRP1* | 9 | 12698611 | G | T | ABSENT |
| *UCHL1* | 4 | 41270049 | G | A | ABSENT |
| *UNC93B1* | 11 | 67759230 | G | T | ABSENT |
| *UROD* | 1 | 45479381 | T | C | ABSENT |
| *UROS* | 10 | 127504753 | G | A | ABSENT |
| *UROS* | 10 | 127496045 | C | T | ABSENT |
| *VPS11* | 11 | 118951867 | A | G | ABSENT |
| *VWF* | 12 | 6167087 | A | C | ABSENT |
| *VWF* | 12 | 6127647 | T | C | ABSENT |
| *WIF1* | 12 | 65460443 | G | C | ABSENT |
| *WWP1* | 8 | 87440033 | G | A | ABSENT |
| *YARS2* | 12 | 32903702 | C | T | ABSENT |

**Supplemental Table 2.** Human genes (n=220) orthologous to the genes with non-human animal missense variants extracted from OMIA and included in this study. See Supplemental Table 1 for details. An asterisk next to a gene symbol indicates that the gene is included in the HRT Atlas v1.0 database list of human housekeeping genes.^1^ OMIM, Online Mendelian Inheritance in Man (<https://www.omim.org/>, accessed 2023).

| **Gene symbol** | **OMIM phenotype** |
| --- | --- |
| *ABCA1* | HDL deficiency, type 2;Tangier disease, Autosomal recessive |
| *ABCA12* | Ichthyosis, congenital, autosomal recessive 4A, Autosomal recessive;Ichthyosis, congenital, autosomal recessive 4B (harlequin), Autosomal recessive |
| *ACSL5* | No Mendelian disease association in OMIM |
| *ACVR1* | Fibrodysplasia ossificans progressiva, Autosomal dominant |
| *ADAMTS10* | Weill-Marchesani syndrome 1, recessive, Autosomal recessive |
| *ADAMTS17* | Weill-Marchesani-like syndrome, Autosomal recessive |
| *ADAMTS2* | Ehlers-Danlos syndrome, type VIIC, Autosomal recessive |
| *ADAMTSL2* | Geleophysic dysplasia 1, Autosomal recessive |
| *ADAMTSL4* | Ectopia lentis et pupillae, Autosomal recessive;Ectopia lentis, isolated, autosomal recessive, Autosomal recessive |
| *AGTPBP1* | No Mendelian disease association in OMIM |
| *AGXT* | Hyperoxaluria, primary, type 1, Autosomal recessive |
| *ALDH5A1* | Succinic semialdehyde dehydrogenase deficiency, Autosomal recessive |
| *ALMS1* | Alstrom syndrome, Autosomal recessive |
| *ALPL* | Hypophosphatasia, adult, Autosomal recessive, Autosomal dominant;Hypophosphatasia, childhood, Autosomal recessive;Hypophosphatasia, infantile, Autosomal recessive;Odontohypophosphatasia, Autosomal recessive, Autosomal dominant |
| *AR* | Androgen insensitivity, X-linked recessive;Androgen insensitivity, partial, with or without breast cancer, X-linked recessive;Hypospadias 1, X-linked, X-linked recessive;Spinal and bulbar muscular atrophy of Kennedy, X-linked recessive |
| *ARSB* | Mucopolysaccharidosis type VI (Maroteaux-Lamy), Autosomal recessive |
| *ARSG* | Usher syndrome, type IV |
| *ASIP* | No Mendelian disease association in OMIM |
| *ASPA* | Canavan disease, Autosomal recessive |
| *ASPRV1* | No Mendelian disease association in OMIM |
| *ATF2* | No Mendelian disease association in OMIM |
| *ATG4D* | No Mendelian disease association in OMIM |
| *ATP13A2* | Kufor-Rakeb syndrome, Autosomal recessive;Spastic paraplegia 78, autosomal recessive, Autosomal recessive |
| *ATP2A1* | Brody myopathy, Autosomal recessive |
| *ATP7A* | Menkes disease, X-linked recessive;Occipital horn syndrome, X-linked recessive;Spinal muscular atrophy, distal, X-linked 3, X-linked recessive |
| *BCKDHA* | Maple syrup urine disease, type Ia, Autosomal recessive |
| *BEST1* | Bestrophinopathy, autosomal recessive;Macular dystrophy, vitelliform, 2, Autosomal dominant;Microcornea, rod-cone dystrophy, cataract, and posterior staphyloma, Autosomal dominant;Retinitis pigmentosa, concentric;Retinitis pigmentosa-50;Vitreoretinochoroidopathy, Autosomal dominant |
| *BMP15* | Ovarian dysgenesis 2;Premature ovarian failure 4 |
| *BMP3* | No Mendelian disease association in OMIM |
| *BMPR1B* | Acromesomelic dysplasia, Demirhan type, Autosomal recessive;Brachydactyly, type A1, D, Autosomal dominant;Brachydactyly, type A2, Autosomal dominant |
| *BRAF* | Cardiofaciocutaneous syndrome, Autosomal dominant;Colorectal cancer, somatic (3);LEOPARD syndrome 3, Autosomal dominant;Melanoma, malignant, somatic (3);Nonsmall cell lung cancer, somatic (3);Noonan syndrome 7, Autosomal dominant |
| *CAD* | Epileptic encephalopathy, early infantile, 50, Autosomal recessive |
| *CAPN1* | Spastic paraplegia 76, autosomal recessive, Autosomal recessive |
| *CAT* | Acatalasemia |
| *CD320 ** | Methylmalonic aciduria, transient, due to transcobalamin receptor defect |
| *CDH23* | Deafness, autosomal recessive 12, Autosomal recessive;Usher syndrome, type 1D, Autosomal recessive, Digenic recessive;Usher syndrome, type 1D/F digenic, Autosomal recessive, Digenic recessive |
| *CHAT* | Myasthenic syndrome, congenital, 6, presynaptic, Autosomal recessive |
| *CHST6* | Macular corneal dystrophy, Autosomal recessive |
| *CLCN1* | Myotonia congenita, dominant, Autosomal dominant;Myotonia congenita, recessive, Autosomal recessive;Myotonia levior, recessive (3) |
| *CLN6* | Ceroid lipofuscinosis, neuronal, 6, Autosomal recessive;Ceroid lipofuscinosis, neuronal, Kufs type, adult onset, Autosomal recessive |
| *CNGA3* | Achromatopsia 2, Autosomal recessive |
| *CNGB3* | Achromatopsia 3, Autosomal recessive;Macular degeneration, juvenile, Autosomal recessive |
| *CNTNAP1* | Lethal congenital contracture syndrome 7, Autosomal recessive |
| *COL10A1* | Metaphyseal chondrodysplasia, Schmid type, Autosomal dominant |
| *COL11A2* | Deafness, autosomal dominant 13, Autosomal dominant;Deafness, autosomal recessive 53, Autosomal recessive;Fibrochondrogenesis 2, Autosomal recessive, Autosomal dominant;Otospondylomegaepiphyseal dysplasia, Autosomal recessive;Stickler syndrome, type III, Autosomal dominant;Weissenbacher-Zweymuller syndrome, Autosomal dominant |
| *COL1A1* | Caffey disease, Autosomal dominant;Ehlers-Danlos syndrome, classic, Autosomal dominant;Ehlers-Danlos syndrome, type VIIA, Autosomal dominant;Osteogenesis imperfecta, type I, Autosomal dominant;Osteogenesis imperfecta, type II, Autosomal dominant;Osteogenesis imperfecta, type III, Autosomal dominant;Osteogenesis imperfecta, type IV, Autosomal dominant |
| *COL2A1* | Achondrogenesis, type II or hypochondrogenesis, Autosomal dominant;Avascular necrosis of the femoral head, Autosomal dominant;Czech dysplasia, Autosomal dominant;Epiphyseal dysplasia, multiple, with myopia and deafness, Autosomal dominant;Kniest dysplasia, Autosomal dominant;Legg-Calve-Perthes disease, Autosomal dominant;Osteoarthritis with mild chondrodysplasia, Autosomal dominant;Otospondylomegaepiphyseal dysplasia, Autosomal recessive;Platyspondylic skeletal dysplasia, Torrance type, Autosomal dominant;SED congenita, Autosomal dominant;SMED Strudwick type, Autosomal dominant;Spondyloepiphyseal dysplasia, Stanescu type, Autosomal dominant;Spondyloperipheral dysplasia, Autosomal dominant;Stickler sydrome, type I, nonsyndromic ocular, Autosomal dominant;Stickler syndrome, type I, Autosomal dominant;Vitreoretinopathy with phalangeal epiphyseal dysplasia (3) |
| *COL5A1* | Ehlers-Danlos syndrome, classic type, Autosomal dominant |
| *COL5A2* | Ehlers-Danlos syndrome, classic type, Autosomal dominant |
| *COL6A3* | Bethlem myopathy 1, Autosomal recessive, Autosomal dominant;Dystonia 27, Autosomal recessive;Ullrich congenital muscular dystrophy 1, Autosomal recessive, Autosomal dominant |
| *COL7A1* | EBD inversa, Autosomal recessive;EBD, Bart type, Autosomal dominant;EBD, localisata variant (3);Epidermolysis bullosa dystrophica, AD, Autosomal dominant;Epidermolysis bullosa dystrophica, AR, Autosomal recessive;Epidermolysis bullosa pruriginosa, Autosomal recessive, Autosomal dominant;Epidermolysis bullosa, pretibial, Autosomal recessive, Autosomal dominant;Toenail dystrophy, isolated, Autosomal dominant;Transient bullous of the newborn, Autosomal recessive, Autosomal dominant |
| *COLQ* | Myasthenic syndrome, congenital, 5, Autosomal recessive |
| *COPA ** | No Mendelian disease association in OMIM |
| *CORIN* | Preeclampsia/eclampsia 5 |
| *CPT1C* | No Mendelian disease association in OMIM |
| *CSNK1G2 ** | No Mendelian disease association in OMIM |
| *CTSD ** | Ceroid lipofuscinosis, neuronal, 10, Autosomal recessive |
| *CYB5R3 ** | Methemoglobinemia, type I, Autosomal recessive;Methemoglobinemia, type II, Autosomal recessive |
| *CYP26C1* | Focal facial dermal dysplasia 4, Autosomal recessive |
| *DKK4* | No Mendelian disease association in OMIM |
| *DMD* | Becker muscular dystrophy, X-linked recessive;Cardiomyopathy, dilated, 3B, X-linked;Duchenne muscular dystrophy, X-linked recessive |
| *DNM1* | Epileptic encephalopathy, early infantile, 31, Autosomal dominant |
| *DNM2* | Charcot-Marie-Tooth disease, axonal, type 2M, Autosomal dominant;Charcot-Marie-Tooth disease, dominant intermediate B, Autosomal dominant;Lethal congenital contracture syndrome 5, Autosomal recessive;Myopathy, centronuclear, Autosomal dominant |
| *DPYS* | Dihydropyrimidinuria, Autosomal recessive |
| *DSP* | Arrhythmogenic right ventricular dysplasia 8, Autosomal dominant;Cardiomyopathy, dilated, with woolly hair and keratoderma, Autosomal recessive;Dilated cardiomyopathy with woolly hair, keratoderma, and tooth agenesis, Autosomal dominant;Epidermolysis bullosa, lethal acantholytic, Autosomal recessive;Keratosis palmoplantaris striata II;Skin fragility-woolly hair syndrome, Autosomal recessive |
| *DUOX2* | Thyroid dyshormonogenesis 6, Autosomal recessive |
| *EDN2* | No Mendelian disease association in OMIM |
| *EDNRA* | Mandibulofacial dysostosis with alopecia, Autosomal dominant |
| *ENAM* | Amelogenesis imperfecta, type IB, Autosomal dominant;Amelogenesis imperfecta, type IC, Autosomal recessive |
| *ENTREP2* | No Mendelian disease association in OMIM |
| *ETFDH* | Glutaric acidemia IIC, Autosomal recessive |
| *F11* | Factor XI deficiency, autosomal dominant;Factor XI deficiency, autosomal recessive |
| *F12* | Angioedema, hereditary, type III, Autosomal dominant;Factor XII deficiency, Autosomal recessive |
| *F8* | Hemophilia A, X-linked recessive |
| *F9* | Hemophilia B, X-linked recessive;Thrombophilia, X-linked, due to factor IX defect |
| *FAM20C* | Raine syndrome, Autosomal recessive |
| *FAM83G* | No Mendelian disease association in OMIM |
| *FBN1* | Acromicric dysplasia, Autosomal dominant;Ectopia lentis, familial, Autosomal dominant;Geleophysic dysplasia 2, Autosomal dominant;MASS syndrome;Marfan lipodystrophy syndrome, Autosomal dominant;Marfan syndrome, Autosomal dominant;Stiff skin syndrome, Autosomal dominant;Weill-Marchesani syndrome 2, dominant, Autosomal dominant |
| *FGF5* | Trichomegaly, Autosomal recessive |
| *FGFR2* | Antley-Bixler syndrome without genital anomalies or disordered steroidogenesis, Autosomal recessive;Apert syndrome, Autosomal dominant;Beare-Stevenson cutis gyrata syndrome, Autosomal dominant;Bent bone dysplasia syndrome, Autosomal dominant;Craniofacial-skeletal-dermatologic dysplasia, Autosomal dominant;Craniosynostosis, nonspecific (3);Crouzon syndrome, Autosomal dominant;Gastric cancer, somatic;Jackson-Weiss syndrome, Autosomal dominant;LADD syndrome, Autosomal dominant;Pfeiffer syndrome, Autosomal dominant;Saethre-Chotzen syndrome, Autosomal dominant;Scaphocephaly and Axenfeld-Rieger anomaly (3);Scaphocephaly, maxillary retrusion, and mental retardation |
| *FGFR3* | Achondroplasia, Autosomal dominant;Bladder cancer, somatic;CATSHL syndrome, Autosomal recessive, Autosomal dominant;Cervical cancer, somatic;Colorectal cancer, somatic;Crouzon syndrome with acanthosis nigricans, Autosomal dominant;Hypochondroplasia, Autosomal dominant;LADD syndrome, Autosomal dominant;Muenke syndrome, Autosomal dominant;Nevus, epidermal, somatic;SADDAN, Autosomal dominant;Spermatocytic seminoma, somatic;Thanatophoric dysplasia, type I, Autosomal dominant;Thanatophoric dysplasia, type II, Autosomal dominant |
| *FLCN ** | Birt-Hogg-Dube syndrome, Autosomal dominant;Colorectal cancer, somatic;Pneumothorax, primary spontaneous, Autosomal dominant;Renal carcinoma, chromophobe, somatic |
| *FZD7* | No Mendelian disease association in OMIM |
| *G6PC1* | Glycogen storage disease Ia, Autosomal recessive |
| *GALC* | Krabbe disease, Autosomal recessive |
| *GART* | No Mendelian disease association in OMIM |
| *GDF9* | No associated disease in OMIM |
| *GFAP* | Alexander disease, Autosomal dominant |
| *GLB1* | GM1-gangliosidosis, type I, Autosomal recessive;GM1-gangliosidosis, type II, Autosomal recessive;GM1-gangliosidosis, type III, Autosomal recessive;Mucopolysaccharidosis type IVB (Morquio), Autosomal recessive |
| *GUSB* | Mucopolysaccharidosis VII, Autosomal recessive |
| *HCRTR2* | No Mendelian disease association in OMIM |
| *HES7* | Spondylocostal dysostosis 4, autosomal recessive, Autosomal recessive |
| *HEXA* | GM2-gangliosidosis, several forms, Autosomal recessive;Tay-Sachs disease, Autosomal recessive |
| *HMBS* | Porphyria, acute intermittent, Autosomal dominant;Porphyria, acute intermittent, nonerythroid variant, Autosomal dominant |
| *IARS1* | No Mendelian disease association in OMIM |
| *IFT122* | Cranioectodermal dysplasia 1, Autosomal recessive |
| *ITGA2B* | Bleeding disorder, platelet-type, 16, autosomal dominant, Autosomal dominant;Glanzmann thrombasthenia, Autosomal recessive;Thrombocytopenia, neonatal alloimmune, BAK antigen related (3) |
| *ITGA3* | Interstitial lung disease, nephrotic syndrome, and epidermolysis bullosa, congenital, Autosomal recessive |
| *ITGB2* | Leukocyte adhesion deficiency, Autosomal recessive |
| *KCNG1* | No Mendelian disease association in OMIM |
| *KCNJ10* | Enlarged vestibular aqueduct, digenic, Autosomal recessive;SESAME syndrome, Autosomal recessive |
| *KDM2B* | No Mendelian disease association in OMIM |
| *KDSR* | Lymphoma/leukemia, B-cell, variant (1) |
| *KIF1C* | Spastic ataxia 2, autosomal recessive, Autosomal recessive |
| *KIF3B* | No Mendelian disease association in OMIM |
| *KIT* | Gastrointestinal stromal tumor, familial, Autosomal dominant, Isolated cases;Germ cell tumors, Somatic mutation;Leukemia, acute myeloid, Autosomal dominant;Mast cell disease, Autosomal dominant;Piebaldism, Autosomal dominant |
| *KLKB1* | Fletcher factor (prekallikrein) deficiency, Autosomal recessive |
| *KRT25* | Woolly hair, autosomal recessive 3, Autosomal recessive |
| *KRT27* | No Mendelian disease association in OMIM |
| *KRT5* | Dowling-Degos disease 1, Autosomal dominant;Epidermolysis bullosa simplex, Dowling-Meara type, Autosomal dominant;Epidermolysis bullosa simplex, Koebner type, Autosomal dominant;Epidermolysis bullosa simplex, Weber-Cockayne type, Autosomal dominant;Epidermolysis bullosa simplex, recessive 1, Autosomal recessive;Epidermolysis bullosa simplex-MP, Autosomal dominant;Epidermylysis bullosa simplex-MCR |
| *KRT71* | No Mendelian disease association in OMIM |
| *L2HGDH* | L-2-hydroxyglutaric aciduria, Autosomal recessive |
| *LAMA3* | Epidermolysis bullosa, generalized atrophic benign, Autosomal recessive;Epidermolysis bullosa, junctional, Herlitz type, Autosomal recessive;Laryngoonychocutaneous syndrome, Autosomal recessive |
| *LAMB3* | Amelogenesis imperfecta, type IA, Autosomal dominant;Epidermolysis bullosa, junctional, Herlitz type, Autosomal recessive;Epidermolysis bullosa, junctional, non-Herlitz type, Autosomal recessive |
| *LAMP3* | No Mendelian disease association in OMIM |
| *LDLR* | Hypercholesterolemia, familial, Autosomal dominant;LDL cholesterol level QTL2, Autosomal dominant |
| *LOXHD1* | Deafness, autosomal recessive 77, Autosomal recessive |
| *LRP4* | Cenani-Lenz syndactyly syndrome, Autosomal recessive;Sclerosteosis 2, Autosomal recessive, Autosomal dominant |
| *LVRN* | No Mendelian disease association in OMIM |
| *LYST* | Chediak-Higashi syndrome, Autosomal recessive |
| *MAN2B1* | Mannosidosis, alpha-, types I and II, Autosomal recessive |
| *MARS2* | Spastic ataxia 3, autosomal recessive, Autosomal recessive |
| *MC1R* | Skin/hair/eye pigmentation |
| *MC4R* | Obesity, autosomal dominant, Autosomal recessive, Autosomal dominant, Multifactorial |
| *MITF* | COMMAD syndrome, Autosomal recessive;Tietz albinism-deafness syndrome, Autosomal dominant;Waardenburg syndrome, type 2A, Autosomal dominant;Waardenburg syndrome/ocular albinism, digenic, Autosomal dominant |
| *MLPH* | Griscelli syndrome, type 3, Autosomal recessive |
| *MOCOS* | Xanthinuria, type II, Autosomal recessive |
| *MRC2* | No Mendelian disease association in OMIM |
| *MSTN* | Muscle hypertrophy |
| *MTBP* | No Mendelian disease association in OMIM |
| *MTM1* | Myotubular myopathy, X-linked, X-linked recessive |
| *MX1* | No Mendelian disease association in OMIM |
| *MYBPC1* | Congenital myopathy 16 |
| *MYBPC3* | Cardiomyopathy, dilated, 1MM, Autosomal dominant;Cardiomyopathy, hypertrophic, 4, Autosomal dominant;Left ventricular noncompaction 10, Autosomal dominant |
| *MYH1* | No Mendelian disease association in OMIM |
| *MYH7* | Cardiomyopathy, dilated, 1S, Autosomal dominant;Cardiomyopathy, hypertrophic, 1, Autosomal dominant;Laing distal myopathy, Autosomal dominant;Left ventricular noncompaction 5, Autosomal dominant;Myopathy, myosin storage, autosomal dominant, Autosomal dominant;Myopathy, myosin storage, autosomal recessive, Autosomal recessive;Scapuloperoneal syndrome, myopathic type, Autosomal dominant |
| *MYO7A* | Deafness, autosomal dominant 11, Autosomal dominant;Deafness, autosomal recessive 2, Autosomal recessive;Usher syndrome, type 1B, Autosomal recessive |
| *NAGLU* | Mucopolysaccharidosis type IIIB (Sanfilippo B), Autosomal recessive |
| *NAPEPLD* | No Mendelian disease association in OMIM |
| *NDRG1* | Charcot-Marie-Tooth disease, type 4D, Autosomal recessive |
| *NECAP1* | No Mendelian disease association in OMIM |
| *NHLRC2* | No Mendelian disease association in OMIM |
| *NPC1* | Niemann-Pick disease, type C1, Autosomal recessive;Niemann-Pick disease, type D, Autosomal recessive |
| *NSDHL* | CHILD syndrome, X-linked dominant;CK syndrome, X-linked recessive |
| *NUBPL* | Mitochondrial complex I deficiency, Autosomal recessive, X-linked dominant, Mitochondrial |
| *P3H2* | Myopia, high, with cataract and vitreoretinal degeneration, Autosomal recessive |
| *PAX3* | Craniofacial-deafness-hand syndrome, Autosomal dominant;Rhabdomyosarcoma 2, alveolar, Autosomal recessive;Waardenburg syndrome, type 1, Autosomal dominant;Waardenburg syndrome, type 3, Autosomal recessive, Autosomal dominant |
| *PCDH15* | Deafness, autosomal recessive 23, Autosomal recessive;Usher syndrome, type 1D/F digenic, Autosomal recessive, Digenic recessive;Usher syndrome, type 1F, Autosomal recessive |
| *PFAS* | No Mendelian disease association in OMIM |
| *PFKM* | Glycogen storage disease VII, Autosomal recessive |
| *PIGN* | Multiple congenital anomalies-hypotonia-seizures syndrome 1, Autosomal recessive |
| *PITX3* | Anterior segment dysgenesis 1, multiple subtypes, Autosomal dominant;Cataract 11, multiple types, Autosomal dominant;Cataract 11, syndromic, Autosomal dominant |
| *PKD1* | Polycystic kidney disease, adult type I, Autosomal dominant |
| *PKLR* | Adenosine triphosphate, elevated, of erythrocytes, Autosomal dominant;Pyruvate kinase deficiency, Autosomal recessive |
| *PLN* | Cardiomyopathy, dilated, 1P;Cardiomyopathy, hypertrophic, 18, Autosomal dominant |
| *PLOD1 ** | Ehlers-Danlos syndrome, type VI, Autosomal recessive |
| *PLP1* | Pelizaeus-Merzbacher disease, X-linked recessive;Spastic paraplegia 2, X-linked, X-linked recessive |
| *PLP2* | No Mendelian disease association in OMIM |
| *PMEL* | No Mendelian disease association in OMIM |
| *PNKP* | Ataxia-oculomotor apraxia 4, Autosomal recessive;Microcephaly, seizures, and developmental delay, Autosomal recessive |
| *PNPLA8* | No Mendelian disease association in OMIM |
| *PPARD* | No Mendelian disease association in OMIM |
| *PPIB ** | Osteogenesis imperfecta, type IX, Autosomal recessive |
| *PRCD* | Retinitis pigmentosa 36 |
| *PRKAG3* | [No associated Mendelian disease in OMIM] Skeletal muscle glycogen content and metabolism trait locus |
| *PRL* | No Mendelian disease association in OMIM |
| *PRNP* | Cerebral amyloid angiopathy, PRNP-related, Autosomal dominant;Creutzfeldt-Jakob disease, Autosomal dominant;Gerstmann-Straussler disease, Autosomal dominant;Huntington disease-like 1, Autosomal dominant;Insomnia, fatal familial, Autosomal dominant;Prion disease with protracted course, Autosomal dominant |
| *PSMB7 ** | No Mendelian disease association in OMIM |
| *PYGM* | McArdle disease, Autosomal recessive |
| *RAB24* | No Mendelian disease association in OMIM |
| *RABGGTB ** | No Mendelian disease association in OMIM |
| *RASGRP2* | No Mendelian disease association in OMIM |
| *RDH5* | Fundus albipunctatus, Autosomal recessive, Autosomal dominant |
| *RETREG1* | Neuropathy, hereditary sensory and autonomic, type IIB, Autosomal recessive |
| *RHO* | Night blindness, congenital stationary, autosomal dominant 1;Retinitis pigmentosa 4, autosomal dominant or recessive, Autosomal recessive, Autosomal dominant;Retinitis punctata albescens, Autosomal recessive, Autosomal dominant |
| *RYR1* | Central core disease, Autosomal recessive, Autosomal dominant;King-Denborough syndrome, Autosomal dominant;Minicore myopathy with external ophthalmoplegia, Autosomal recessive;Neuromuscular disease, congenital, with uniform type 1 fiber, Autosomal recessive, Autosomal dominant |
| *SCN8A* | Epileptic encephalopathy, early infantile, 13, Autosomal dominant;Seizures, benign familial infantile, 5, Autosomal dominant |
| *SEL1L ** | No Mendelian disease association in OMIM |
| *SERPINH1* | Osteogenesis imperfecta, type X, Autosomal recessive |
| *SGSH* | Mucopolysaccharidosis type IIIA (Sanfilippo A), Autosomal recessive |
| *SLC12A1* | Bartter syndrome, type 1, Autosomal recessive |
| *SLC24A5* | Albinism, oculocutaneous, type VI, Autosomal recessive |
| *SLC25A12* | Epileptic encephalopathy, early infantile, 39, Autosomal recessive |
| *SLC25A46* | Neuropathy, hereditary motor and sensory, type VIB, Autosomal recessive |
| *SLC2A9* | Hypouricemia, renal, 2, Autosomal recessive, Autosomal dominant |
| *SLC35A3* | No Mendelian disease association in OMIM |
| *SLC36A1* | No Mendelian disease association in OMIM |
| *SLC39A4* | Acrodermatitis enteropathica, Autosomal recessive |
| *SLC3A1* | Cystinuria, Autosomal recessive, Autosomal dominant |
| *SLC45A2* | Albinism, oculocutaneous, type IV |
| *SLC5A3* | No Mendelian disease association in OMIM |
| *SLC6A5* | Hyperekplexia 3, Autosomal recessive, Autosomal dominant |
| *SLC7A9* | Cystinuria, Autosomal recessive, Autosomal dominant |
| *SMC2* | No Mendelian disease association in OMIM |
| *SOD1 ** | Amyotrophic lateral sclerosis 1, Autosomal recessive, Autosomal dominant |
| *SOWAHB* | No Mendelian disease association in OMIM |
| *SPAST* | Spastic paraplegia 4, autosomal dominant, Autosomal dominant |
| *STAT5B ** | Growth hormone insensitivity with immunodeficiency;Leukemia, acute promyelocytic, somatic |
| *SUGT1 ** | No Mendelian disease association in OMIM |
| *SUV39H2* | No Mendelian disease association in OMIM |
| *TBXT* | No Mendelian disease association in OMIM |
| *TECPR2* | Spastic paraplegia 49, autosomal recessive, Autosomal recessive |
| *TNXB* | Ehlers-Danlos syndrome due to tenascin X deficiency, Autosomal recessive;Vesicoureteral reflux 8, Autosomal dominant |
| *TPO* | Thyroid dyshormonogenesis 2A, Autosomal recessive |
| *TRPV4* | Brachyolmia type 3, Autosomal dominant;Digital arthropathy-brachydactyly, familial, Autosomal dominant;Hereditary motor and sensory neuropathy, type IIc, Autosomal dominant;Metatropic dysplasia, Autosomal dominant;Parastremmatic dwarfism, Autosomal dominant;SED, Maroteaux type, Autosomal dominant;Scapuloperoneal spinal muscular atrophy, Autosomal dominant;Spinal muscular atrophy, distal, congenital nonprogressive, Autosomal dominant;Spondylometaphyseal dysplasia, Kozlowski type, Autosomal dominant |
| *TSEN54* | Pontocerebellar hypoplasia type 2A, Autosomal recessive;Pontocerebellar hypoplasia type 4, Autosomal recessive |
| *TUBB1* | Macrothrombocytopenia, autosomal dominant, TUBB1-related, Autosomal dominant |
| *TUBD1* | No Mendelian disease association in OMIM |
| *TUBGCP5* | No Mendelian disease association in OMIM |
| *TYR* | Albinism, oculocutaneous, type IA, Autosomal recessive;Albinism, oculocutaneous, type IB;Waardenburg syndrome/albinism, digenic, Autosomal dominant |
| *TYRP1* | Albinism, oculocutaneous, type III, Autosomal recessive |
| *UCHL1* | Spastic paraplegia 79, autosomal recessive, Autosomal recessive |
| *UNC93B1* | No Mendelian disease association in OMIM |
| *UROD ** | Porphyria cutanea tarda, Autosomal dominant;Porphyria, hepatoerythropoietic, Autosomal dominant |
| *UROS* | Porphyria, congenital erythropoietic, Autosomal recessive |
| *VPS11* | Leukodystrophy, hypomyelinating, 12, Autosomal recessive |
| *VWF* | von Willebrand disease, type 1, Autosomal dominant;von Willebrand disease, types 2A, 2B, 2M, and 2N, Autosomal recessive, Autosomal dominant;von Willibrand disease, type 3, Autosomal recessive |
| *WIF1* | No Mendelian disease association in OMIM |
| *WWP1* | No Mendelian disease association in OMIM |
| *YARS2* | Myopathy, lactic acidosis, and sideroblastic anemia 2, Autosomal recessive |

**Supplemental Figure 1.** Human karyogram indicating Mendelian diseases and the associated genes, where orthologous variants were observed in both non-human animals (OMIA) and humans (ClinVar). See text for details. Positions are approximate. Created with [BioRender.com](https://biorender.com/).

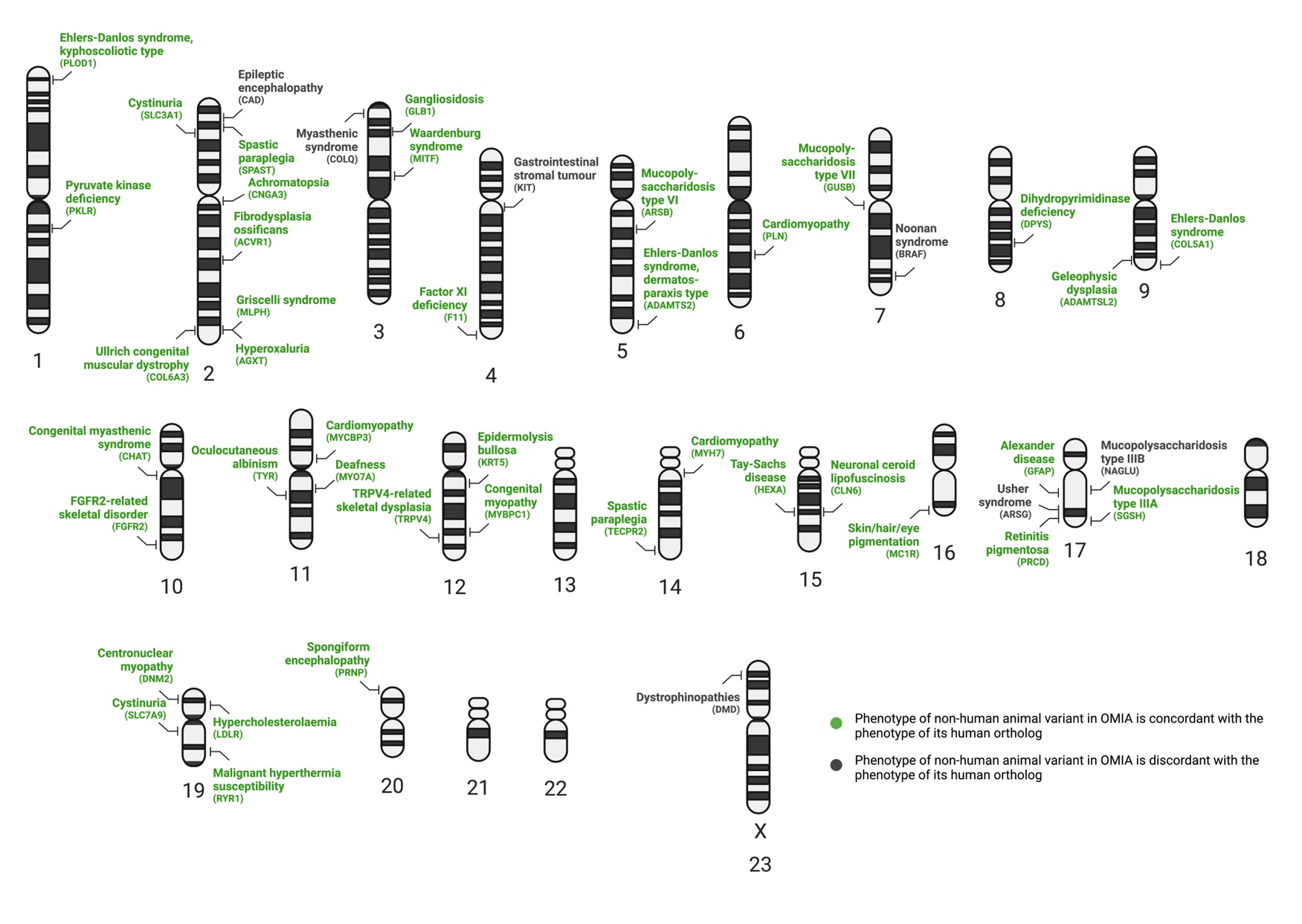

**Supplemental Figure 2.** An example of orthologous germline missense variants in *DNM2* (NM_001005361.3:c.1393C>T) affecting the conserved wild-type arginine residue [p.(Arg465Trp)] and causing centronuclear myopathy in both dogs and humans.^2 3^ Created with [BioRender.com](https://biorender.com/).

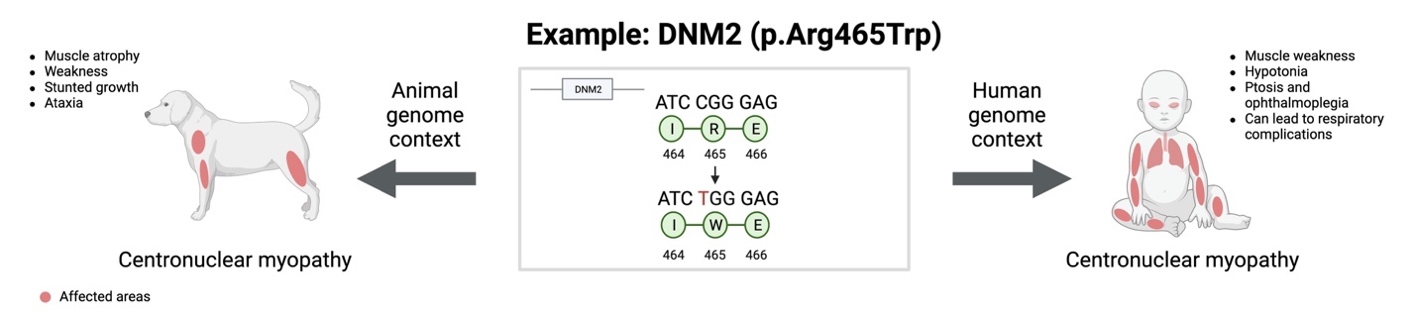

**Supplemental Figure 3.** Odds of the missense variants in non-human animal species extracted from OMIA for which orthologous human variants appear in ClinVar to have been classified in the latter as pathogenic/likely pathogenic (P/LP), compared to benign/likely benign/variant of uncertain significance/conflicting interpretations of pathogenicity (B/LB/VUS/CIP). Created with [BioRender.com](https://biorender.com/).

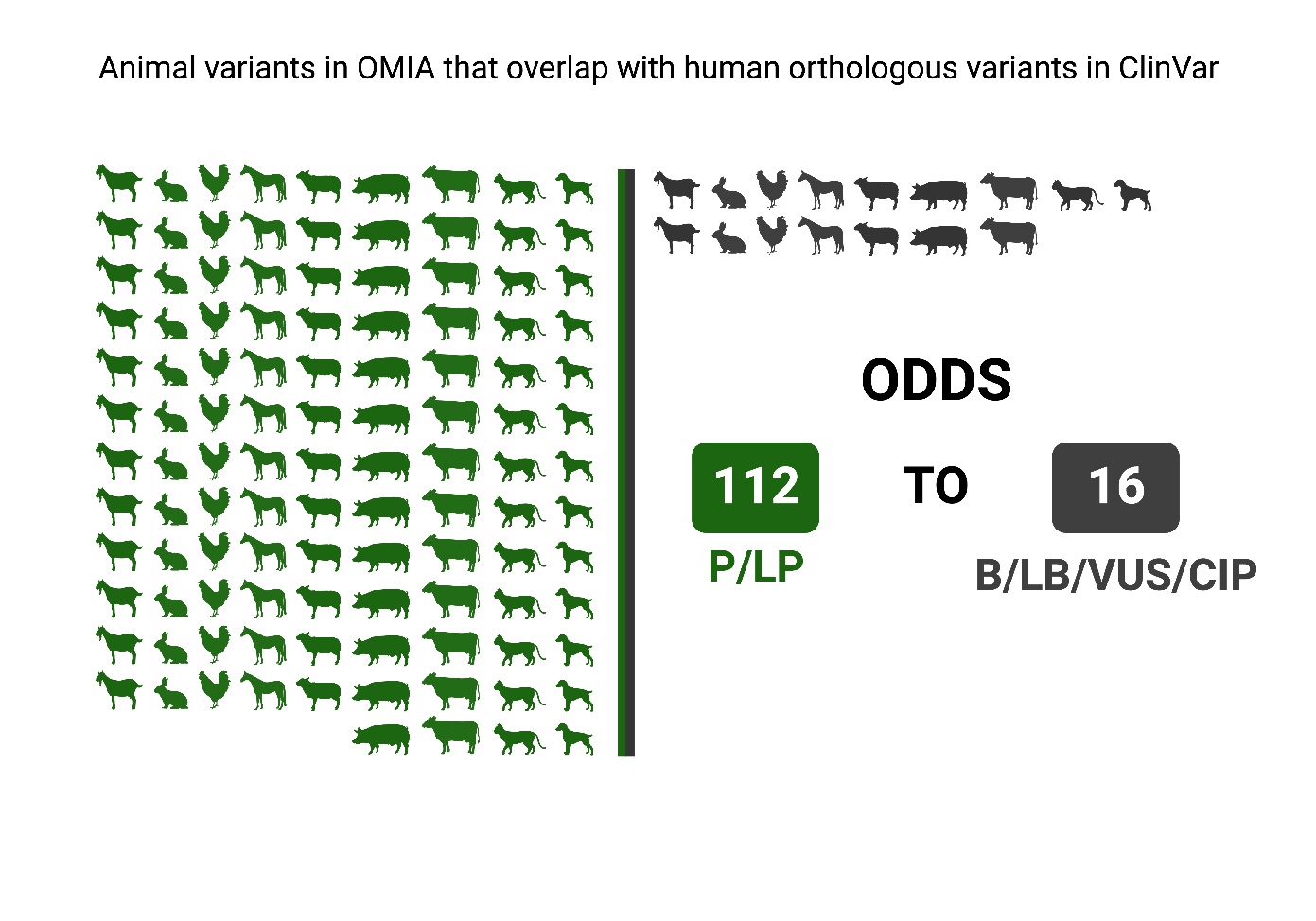

**Supplemental Figure 4.** Bar chart summarizing the evidence type(s) used in publications describing the initial association between a specific missense variant and a phenotype in a non-human animal species, for all variants in OMIA for which orthologous human variants were classified in ClinVar (n=56). Figure created with GraphPad Software.

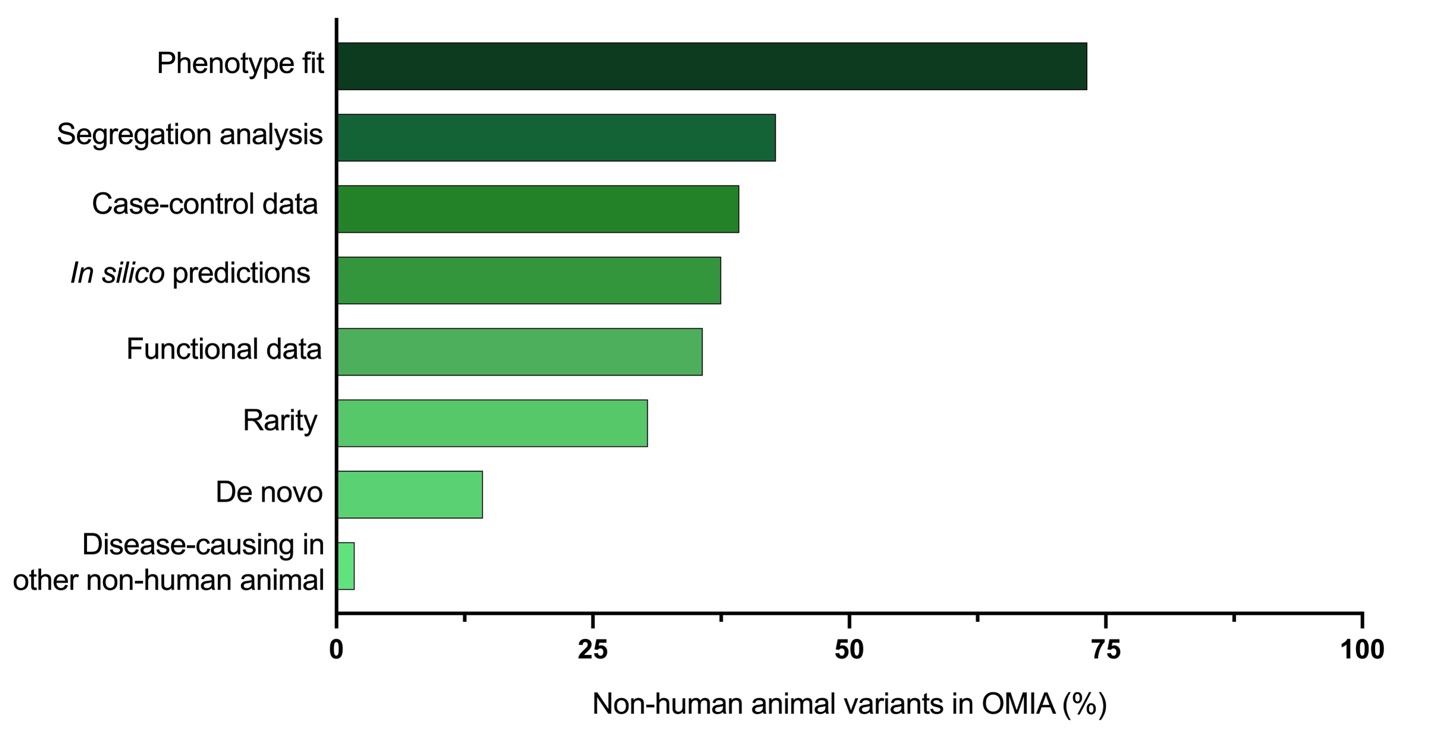

**Supplemental Figure 5.** Boxplots of AlphaMissense^4^ pathogenicity scores for the OMIA missense variants absent from ClinVar, comparing with other missense variation in ClinVar in the same gene set. B/LB, Benign/Likely benign; P/LP, Pathogenic/Likely pathogenic; VUS/CIP, variant of uncertain significance/conflicting interpretations of pathogenicity.

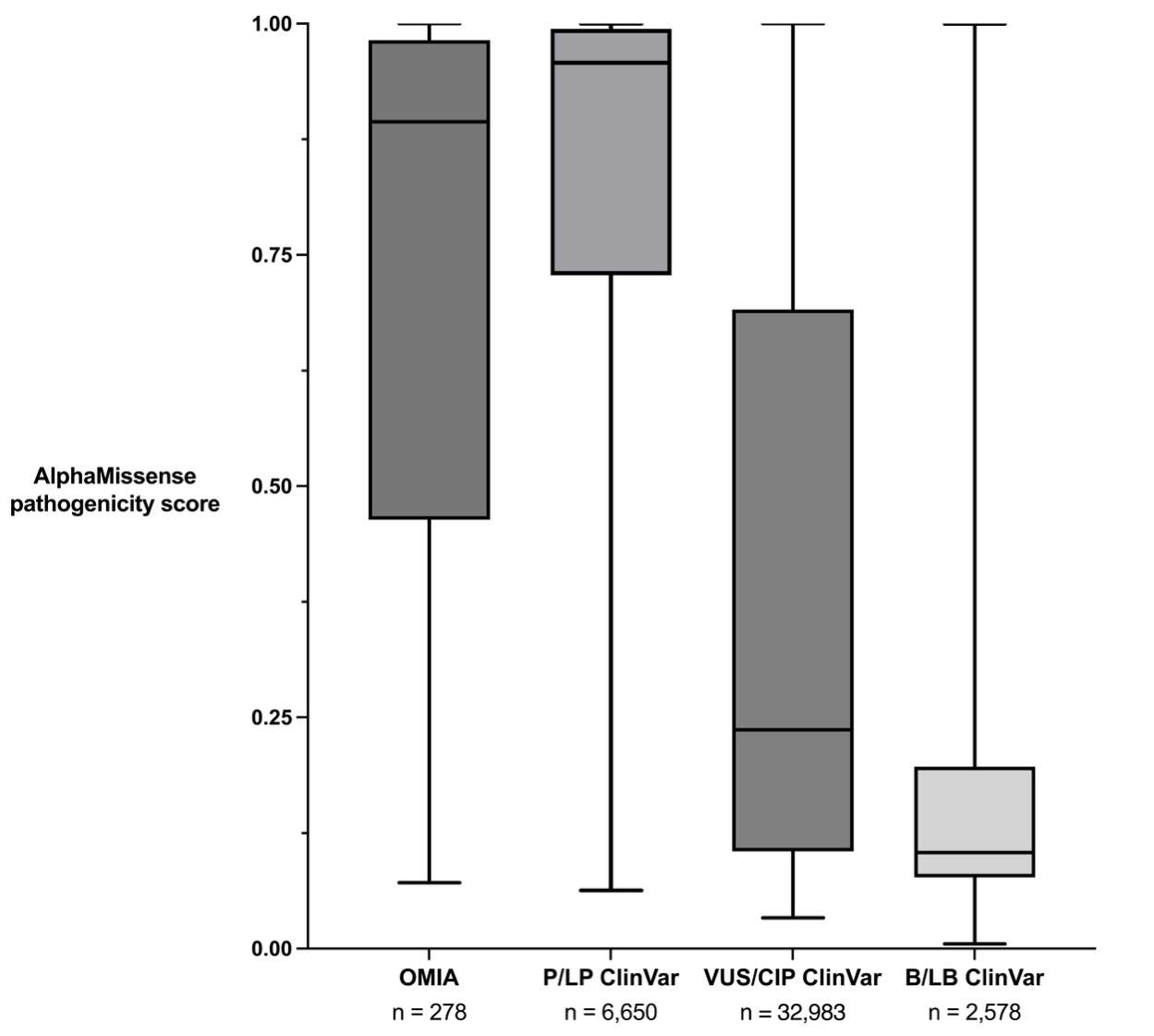
